# Increased expression of ACE2, the SARS-CoV-2 entry receptor, in alveolar and bronchial epithelium of smokers and COPD subjects

**DOI:** 10.1101/2020.05.27.20114298

**Authors:** Merel Jacobs, Hannelore P Van Eeckhoutte, Sara RA Wijnant, Wim Janssens, Guy F Joos, Guy G Brusselle, Ken R Bracke

## Abstract

**Rationale:** Smokers and patients with chronic obstructive pulmonary disease (COPD) are at increased risk for severe Coronavirus Disease 2019 (COVID-19).

**Objectives:** We investigated the expression of the severe acute respiratory syndrome coronavirus 2 (SARS-CoV-2) entry receptor ACE2 and the protease TMPRSS2 in lung tissue from never smokers and smokers with and without COPD.

**Methods:** In a cross-sectional, observational study we measured mRNA expression of ACE2 and TMPRSS2 by RT-PCR in lung tissue samples from 120 well phenotyped subjects. Next, protein levels of ACE2 were visualized by immunohistochemistry on paraffin sections from 87 subjects and quantified in alveolar and bronchial epithelium. Finally, primary human bronchial epithelial cells (HBECs) were cultured at air liquid interface and exposed to air or cigarette smoke.

**Results:** ACE2 mRNA expression was significantly higher in lung tissue from current smokers and subjects with moderate to very severe COPD and correlated with physiological parameters of airway obstruction and emphysema. Pulmonary expression levels of TMPRSS2 were significantly higher in patients with (very) severe COPD and correlated significantly with ACE2 expression. Importantly, protein levels of ACE2 were elevated in both alveolar and bronchial epithelium of current smokers and subjects with moderate to very severe COPD. Finally, TMPRSS2 mRNA expression increased in *in vitro* cultured HBECs upon acute exposure to cigarette smoke.

**Conclusions:** We demonstrate increased expression of ACE2 in lungs of smokers and COPD subjects, which might facilitate host cell entry of SARS-CoV-2. These findings help identifying populations at risk for severe COVID-19.

## INTRODUCTION

Coronavirus Disease 19 (COVID-19) is a novel emerging respiratory disease caused by the severe acute respiratory syndrome coronavirus 2 (SARS-CoV-2), and is causing a vast pandemic with huge medical, social, personal and financial impact. The virus first appeared in Wuhan, China and rapidly spread across the rest of the world, making COVID-19 the third large-scale pandemic caused by coronaviruses after SARS in 2002 and Middle East respiratory syndrome (MERS) in 2012 [1, 2]. The clinical course of the disease ranges from an asymptomatic course to progressive respiratory failure with need for ventilatory support and even death [1].

Angiotensin-converting enzyme 2 (ACE2) was identified as the cell entry receptor used by SARS-CoV-1 and SARS-CoV-2 [1, 3]. ACE2 is a metallopeptidase I and a homologue of the ACE-receptor. Although both receptors play important roles in the renin-angiotensin-aldosterone system (RAAS), they have profoundly different roles. Whereas ACE converts angiotensin I (Ang I) to angiotensin II (Ang II), a potent vasoconstrictor, ACE2 converts both Ang I and Ang II to Ang-(1-9) and Ang-(1-7) respectively, thereby counteracting the effects of ACE and Ang II [4, 5]. Expression of ACE2 was shown in various cells and tissues, including alveolar type II cells and bronchial epithelial cells, as well as in the oral cavity, oesophagus, heart, kidney, and ileum [6]. In addition, transmembrane protease, serine 2 (TMPRSS2) was identified as the host cell protease used by SARS-CoV-2 for S protein priming, which is an essential part of the viral entry process [7].

Although SARS-CoV-2 can infect any individual, most severe cases are reported in those with comorbidities such as hypertension, diabetes, and chronic obstructive pulmonary disease (COPD) [2, 8-10]. COPD is a highly prevalent chronic respiratory disease, characterized by an abnormal inflammatory response to cigarette smoke [11]. Importantly, both smokers and patients with COPD are at increased risk for severe complications and a higher mortality upon SARS-CoV-2 infection [12, 13]. Whether this is due to an increased expression of ACE2 in the lungs, and more specifically in the alveolar tissue where pneumonia occurs in severe COVID-19, needs thorough investigation. A recent paper reported increased ACE2 expression in bronchial brushings and airway epithelia of current smokers and patients with COPD, compared to healthy controls [14]. Previous work done by our group, showed upregulation of dipeptidyl peptidase 4 (DPP4), the viral entry receptor for the MERS coronavirus, in alveolar tissue of smokers and patients with COPD [15].

We hypothesized that the increased risk for a more severe course of COVID-19 in smokers and patients with COPD is due to an increased expression of the SARS-CoV-2 entry receptor ACE2 in the lung. Therefore, we aimed to investigate the expression of ACE2, as well as the host cell protease TMPRSS2, in a large number of lung tissue specimens of well phenotyped subjects, including never smokers and smokers with and without airflow limitation. We quantified ACE2 and TMPRSS2 mRNA expression in the lung, as well as ACE2 protein levels in both alveolar and bronchial epithelium. Next, we investigated the independent determinants of pulmonary ACE2 expression by linear regression analysis. Finally, we determined mRNA expression of ACE2 and TMPRSS2 in cigarette smoke-exposed primary human bronchial epithelial cells.

## METHODS

Methodological details are available in the online data supplement. Study design, hypothesis, methods, analyses, and findings are reported according to the STROBE checklist for cross-sectional studies [16, 17].

### Human lung tissue samples

In this cross-sectional, observational study, we analyzed lung tissue specimens from a total of 134 subjects: 106 lung samples from our large lung tissue biobank at Ghent University Hospital (collected from lung resections for solitary pulmonary tumors and currently encompassing 360 subjects) and 28 samples from explant lungs from end-stage COPD patients collected at UZ Gasthuisberg Leuven, Belgium. All lung samples were collected between January 2002 and February 2020, before the first reported case of COVID-19 in Belgium. A flow-chart illustrating the selection of lung samples for RT-PCR (n=120) and immunohistochemical (n=87) analyses is shown in **Figure E1**. Based on medical history, smoking history, questionnaires and preoperative spirometry, patients were categorized as never-smokers with normal lung function, smokers without airflow limitation or subjects with COPD. Subjects were considered exsmokers when they had quitted smoking for more than 1 year. COPD severity was defined according to the Global Initiative for Chronic Obstructive Lung Disease (GOLD) classification. None of the patients were treated with neo-adjuvant chemotherapy. Lung tissue of patients diagnosed with solitary pulmonary tumors was obtained at a maximum distance from the pulmonary lesions and without signs of retro-obstructive pneumonia or tumor invasion and collected by a pathologist. Written informed consent was obtained from all subjects. This study was approved by the medical ethical committees of the Ghent University Hospital (2011/0114; 2016/0132; 2019/0537) and the University Hospital Gasthuisberg Leuven (S51577).

### Culture of human bronchial epithelial cells

Primary human bronchial epithelial cells (HBECs) were isolated by enzymatic digestion from lung resection specimens derived from 6 donors (1 non-COPD and 5 COPD GOLD stage II or III) during surgery for lung cancer, as described previously [18]. After a 14-day ALI culture period, cells were exposed to mainstream cigarette smoke or air, as described previously, and harvested after 3, 6 or 24 hours [19].

### RNA extraction and real-time PCR-analysis

RNA extraction from lung tissue blocks of 120 subjects (including 28 never-smokers, 36 smokers without airflow limitation, 42 patients with COPD GOLD II and 14 patients with COPD GOLD III-IV, see **Table 1** for patient characteristics), as well as from the ALI cultured HBECs, was performed with the miRNeasy Mini kit (Qiagen, Hilden, Germany), between 2015 and 2020. Next, cDNA was prepared with the EvoScript Universal cDNA Master Kit (Roche) in March 2020, followed by RT-PCR analysis for ACE2, TMPRSS2 and 3 reference genes as described previously [20, 21].

**TABLE 1.**
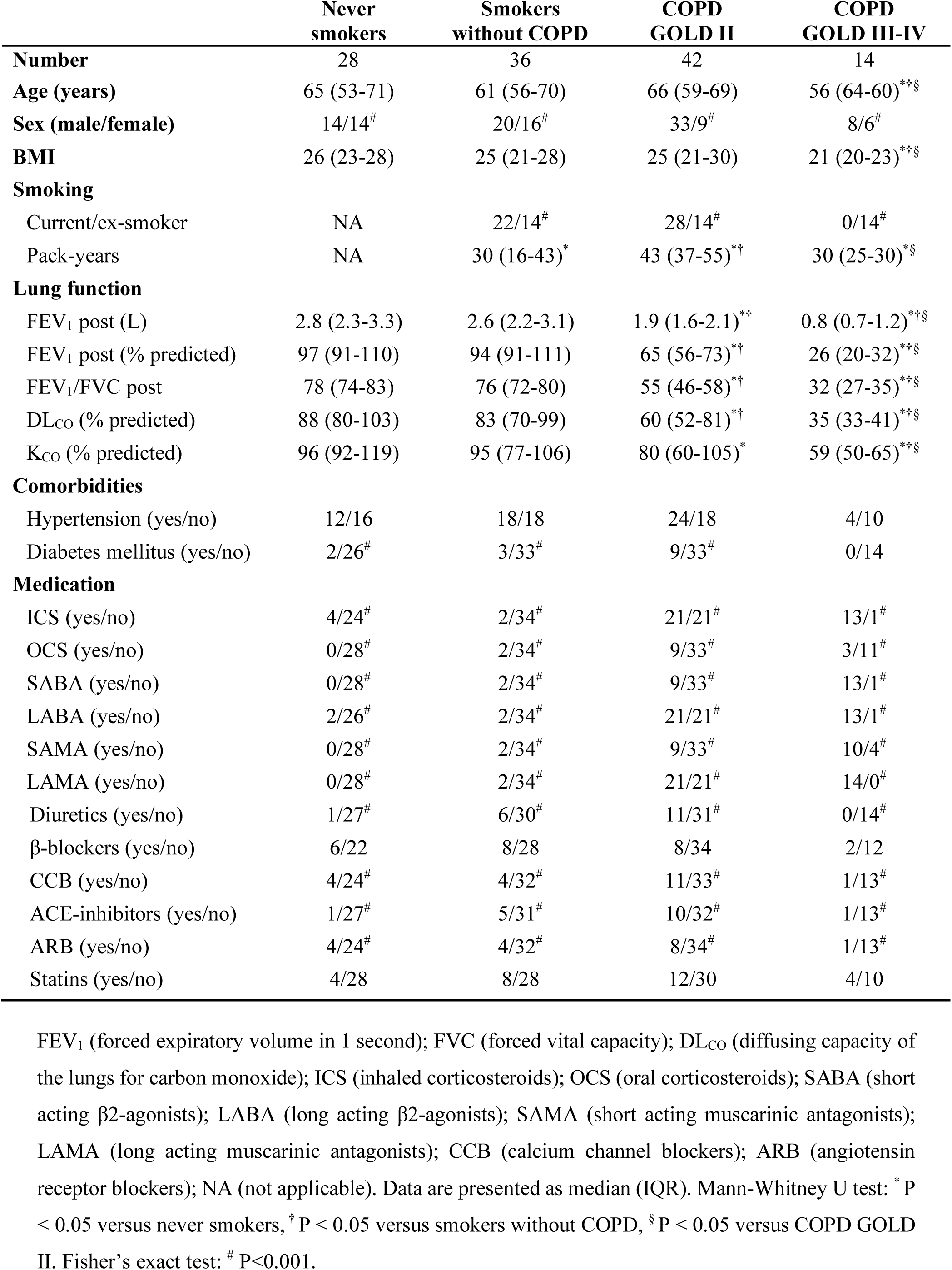
Patient characteristics for mRNA analysis (by qRT-PCR) (n = 120)

### ACE2 immunohistochemistry

Sections from formalin fixed paraffin embedded lung tissue blocks of 87 subjects (including 20 never-smokers, 24 smokers without COPD, 29 subjects with COPD GOLD II and 14 subjects with COPD GOLD III-IV, see supplementary **Table E1** for patient characteristics) were stained for ACE2 in March and April 2020. After antigen retrieval with citrate buffer (Scytek), the slides were incubated with anti-ACE2 antibody (polyclonal rabbit-anti-human, Abcam ab15248). Next, slides were colored with diaminobenzidine (Dako, Carpinteria, CA, USA) and counterstained with Mayer’s hematoxylin (Sigma-Aldrich, St-Louis, MO, USA). The isotype control was rabbit IgG (R&D systems, AB-150-C).

### Quantification of ACE2 in alveolar tissue and airway epithelium

Quantitative measurements of the ACE2-positive signal in alveolar tissue and bronchial epithelium was performed on images of stained paraffin sections as described previously [15]. A detailed description of the methods can be found in the online supplement. Briefly, to quantify the amount of ACE2-positive signal in alveolar tissue, 10 images of alveolar tissue (not containing airways or blood vessels) were recorded from an average of 3 tissue blocks per patient. The area of brown ACE2-positive staining was measured and normalized to the total area of alveolar tissue present in each image. Additionally, the number of ACE2-positive cells was manually counted in each image and again normalized to the total area of alveolar tissue. To quantify the amount of ACE2-positive signal in airway epithelium, images of airways were recorded from an average of 3 tissue blocks per patient. The amount of ACE2-positive signal was measured only in the airway epithelial layer and normalized to the length of the basement membrane (Pbm). The number of airways per patient was between 3 and 20.

### Statistical analysis

Statistical analysis was performed with Sigma Stat software (SPSS 26.0, Chicago, IL, USA) and R3.5.1, using Student’s t-test, Kruskal-Wallis and Mann-Whitney U test, Fisher’s exact test, Spearman’s Rank correlations, and multivariate linear regression analyses. Additional details and covariates for the regression analyses are described in the online supplement. Characteristics of the study population are presented as median (interquartile range). Differences at p-values < 0.05 were considered to be significant (*P<0.05, **P<0.01 and *** P<0.001).

## RESULTS

### ACE2 mRNA expression is increased in lung tissue of smokers and COPD subjects

Using real time quantitative PCR, ACE2 mRNA levels were determined in lung tissue from 120 subjects. Lung tissue from 20 never smokers, 36 smokers without airflow limitation (including 22 current smokers and 14 ex-smokers) and 42 patients with COPD GOLD stage II (including 28 current smokers and 14 ex-smokers) was derived from lobectomy specimens, whereas tissue from 14 subjects with COPD GOLD stage III-IV was derived from explant lungs after lung transplantation. Demographic and clinical patient characteristics are described in **Table 1**.

ACE2 mRNA expression was significantly higher in lung tissue of current smokers without airflow limitation, current smokers with COPD GOLD stage II, and smokers with COPD GOLD stage III-IV, compared to never smokers **(Figure 1A)**. In addition, ex-smokers without airflow limitation showed significantly lower ACE2 mRNA levels, compared to current smokers.

**Figure 1.**
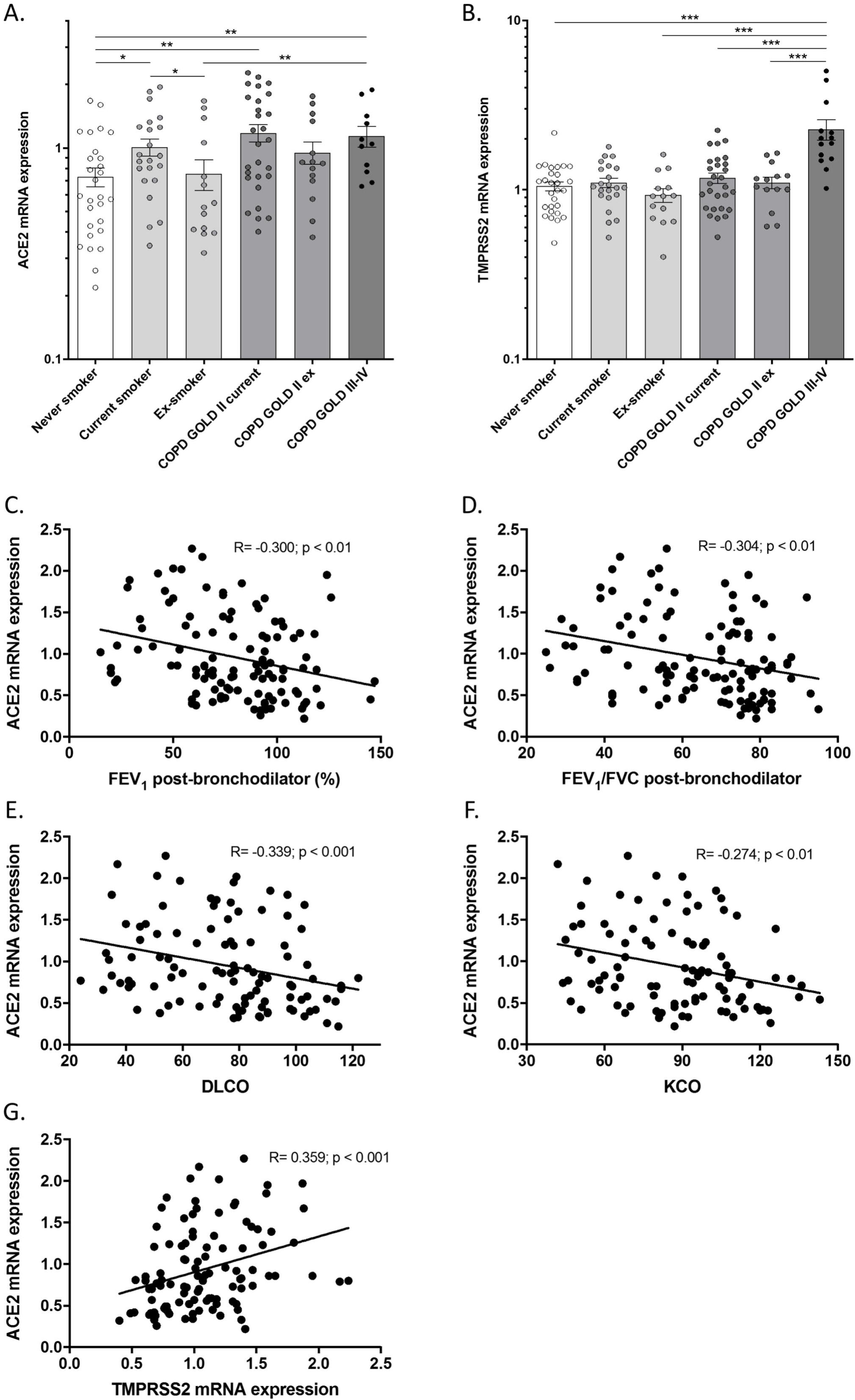
ACE2 mRNA expression is increased in lung tissue from smokers and COPD subjects and correlates with lung function impairment. **(A)** Angiotensin-converting Enzyme-2 (ACE2) and **(B)** Transmembrane protease, serine 2 (TMPRSS2) mRNA expression in lung tissue from never smokers, current and ex-smokers without airflow limitation and current and ex-smokers with mild (GOLD II) or moderate-to-severe (GOLD III-IV) COPD, normalized to the expression of the housekeeping controls glyceraldehyde-3-phosphate dehydrogenase (GAPDH), peptidylprolyl isomerase A (PPIA) and succinate dehydrogenase complex flavoprotein subunit A (SDHA). Data are presented as means ± SEM. Correlations of ACE2 mRNA expression with **(C)** post-bronchodilator forced expiratory volume in 1 second (FEV_1_) % predicted, **(D)** FEV_1_/forced vital capacity (FVC) ratio, **(E)** diffusing capacity of the lungs for carbon monoxide (DL_CO_) % predicted, **(F)** the diffusion coefficient (K_CO_) % predicted and **(G)** TMPRSS2 mRNA expression. Associations for each comparison are expressed as Spearman’s rank correlation coefficient. *P<0.05; **P<0.01; ***P<0.001.

Furthermore, pulmonary ACE2 mRNA expression was inversely correlated with the severity of airflow limitation (post-bronchodilator forced expiratory volume in 1 second (FEV1) % predicted and the ratio of FEV1 to the forced vital capacity (FVC)) and with the diffusing capacity of the lung for carbon monoxide (DL_co_ and K_co_), markers of emphysema **(Figure 1C-F)**.

TMPRSS2 mRNA expression was significantly higher in lung tissue of patients with COPD GOLD stage III-IV, compared to never smokers, smokers without airflow limitation and patients with COPD GOLD stage II **(Figure 1B)**. Moreover, there was a significant positive correlation between pulmonary TMRPSS2 mRNA expression and ACE2 mRNA expression, even after excluding the high TMPRSS2 expressing GOLD stage III-IV patients **(Figure 1G)**.

### ACE2 protein levels are increased in alveolar tissue and bronchial epithelium of smokers and COPD subjects

By immunohistochemical (IHC) staining, ACE2 protein levels were assessed in lung tissue from 87 subjects: 20 never smokers, 24 smokers without COPD (15 current smokers and 9 ex-smokers), 29 patients with COPD GOLD stage II (20 current smokers and 9 ex-smokers) and 14 smokers with COPD GOLD stage III-IV. Patient characteristics are described in supplementary **Table E1**.

ACE2 IHC revealed positive staining in both bronchial and alveolar epithelial cells **(Figure 2)**, with the latter predominantly in alveolar type II cells.

**Figure 2.**
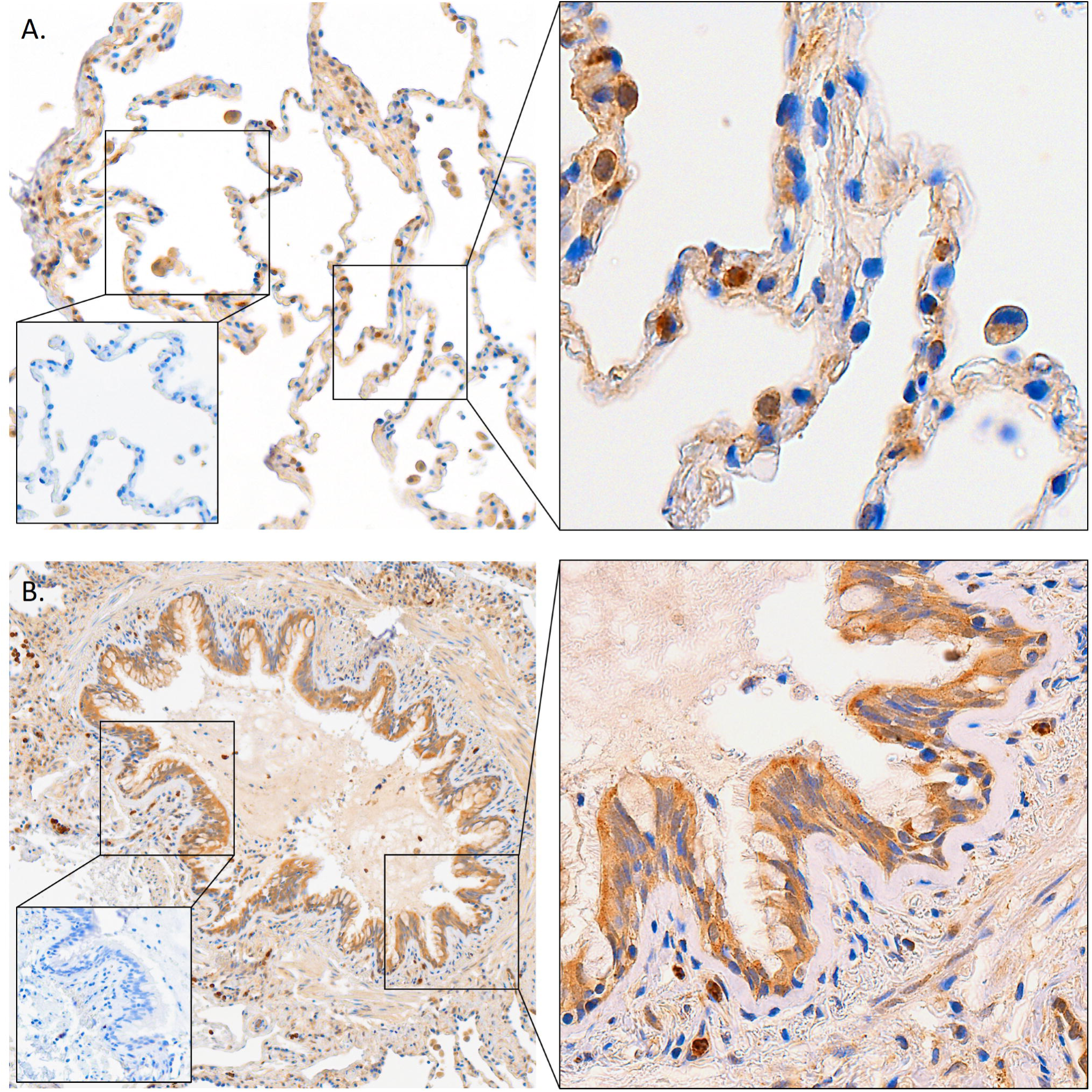
ACE2 immunohistochemistry reveals expression in alveolar and bronchial epithelium. Representative images of ACE2 immunohistochemical staining, revealing positive signal in **(A)** alveolar and **(B)** bronchial epithelium, at a 200x magnification. Small inlays are representative of the negative isotype control staining. Images of a selected area are taken at 400x magnification. ACE2 expression in alveolar epithelium was mainly observed in alveolar type II cells.

Quantification of ACE2 protein levels in alveolar tissue revealed a significantly higher percentage of ACE2-positive alveolar tissue in current smokers without airflow limitation, current smokers with COPD GOLD II, and patients with COPD GOLD III-IV, compared to never smokers **(Figure 3A-E)**. Moreover, the percentage of ACE2-positive alveolar tissue was significantly higher in patients with COPD GOLD III-IV, compared to ex-smokers without airflow limitation and ex-smokers with COPD GOLD II **(Figure 3E)**. Additionally, quantification of the number of ACE2 positive cells in the alveolar epithelium showed a similar increase in current smokers and subjects with COPD (GOLD stages II and III-IV), which reached significance only in (very) severe COPD patients **(Figure E2)**.

**Figure 3.**
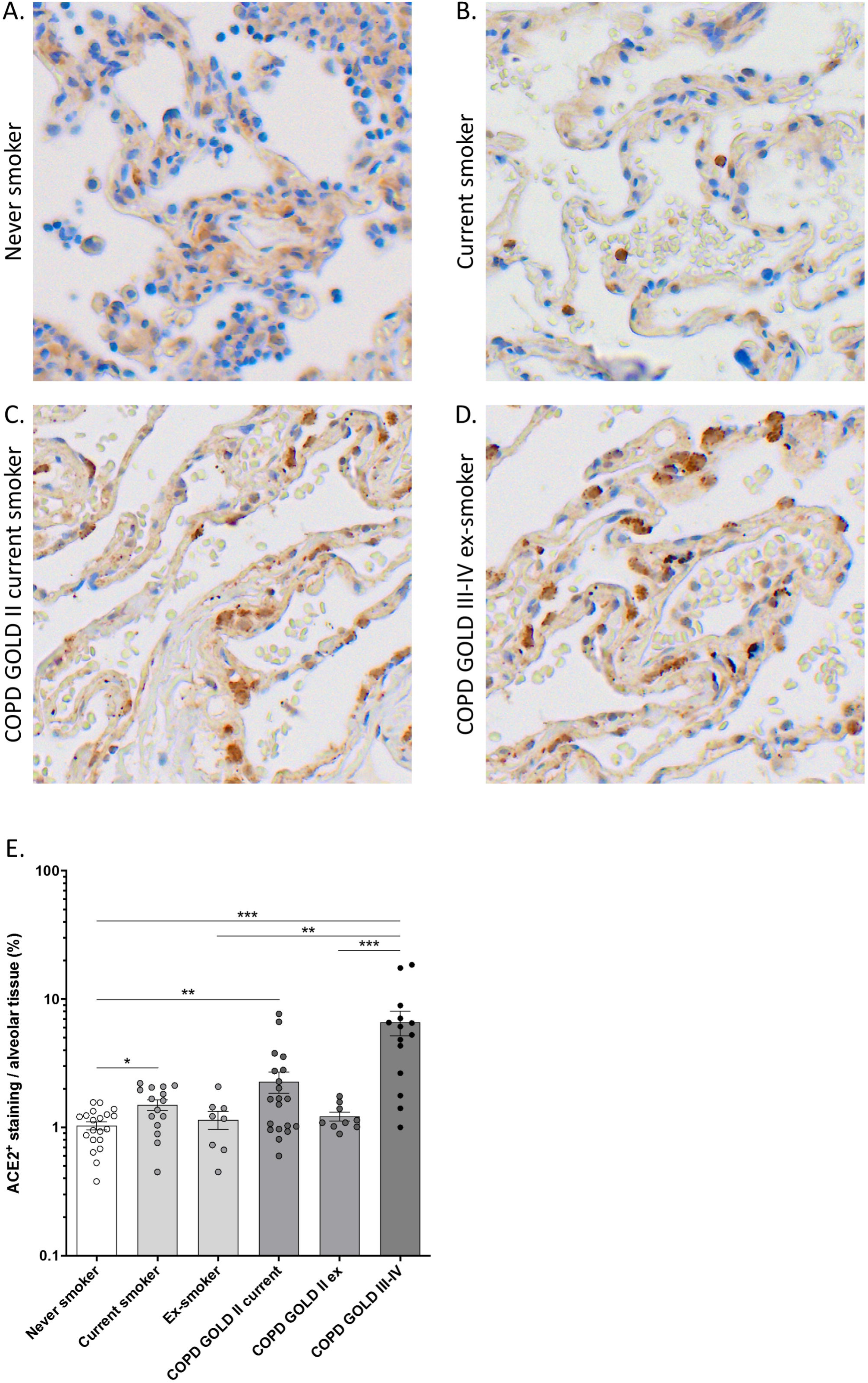
ACE2 protein levels are increased in alveolar tissue from smokers and COPD subjects. Representative images of ACE2 immunohistochemical staining in alveolar tissue from **(A)** never smokers, **(B)** current smokers without airflow limitation, **(C)** current smokers with COPD GOLD stage II and **(D)** ex-smokers with COPD GOLD stage III-IV. **(E)** Quantification of ACE2-positive alveolar tissue using AxioVision software (Zeiss). The area of ACE2-positive signal was normalized to the total area of alveolar tissue present in each analyzed image. Data are presented as means ± SEM. *P<0.05; **P<0.01; ***P<0.001.

Quantification of ACE2 staining in bronchial epithelium revealed numerically higher levels in current smokers without airflow limitation and current smokers with COPD GOLD II, and significantly higher levels in patients with COPD GOLD III-IV, compared to never smokers **(Figure 4A-E)**. Moreover, ACE2 protein levels in bronchial epithelium were significantly higher in patients with COPD GOLD III-IV, compared to ex-smokers without airflow limitation and ex-smokers with COPD GOLD II **(Figure 4E)**.

**Figure 4.**
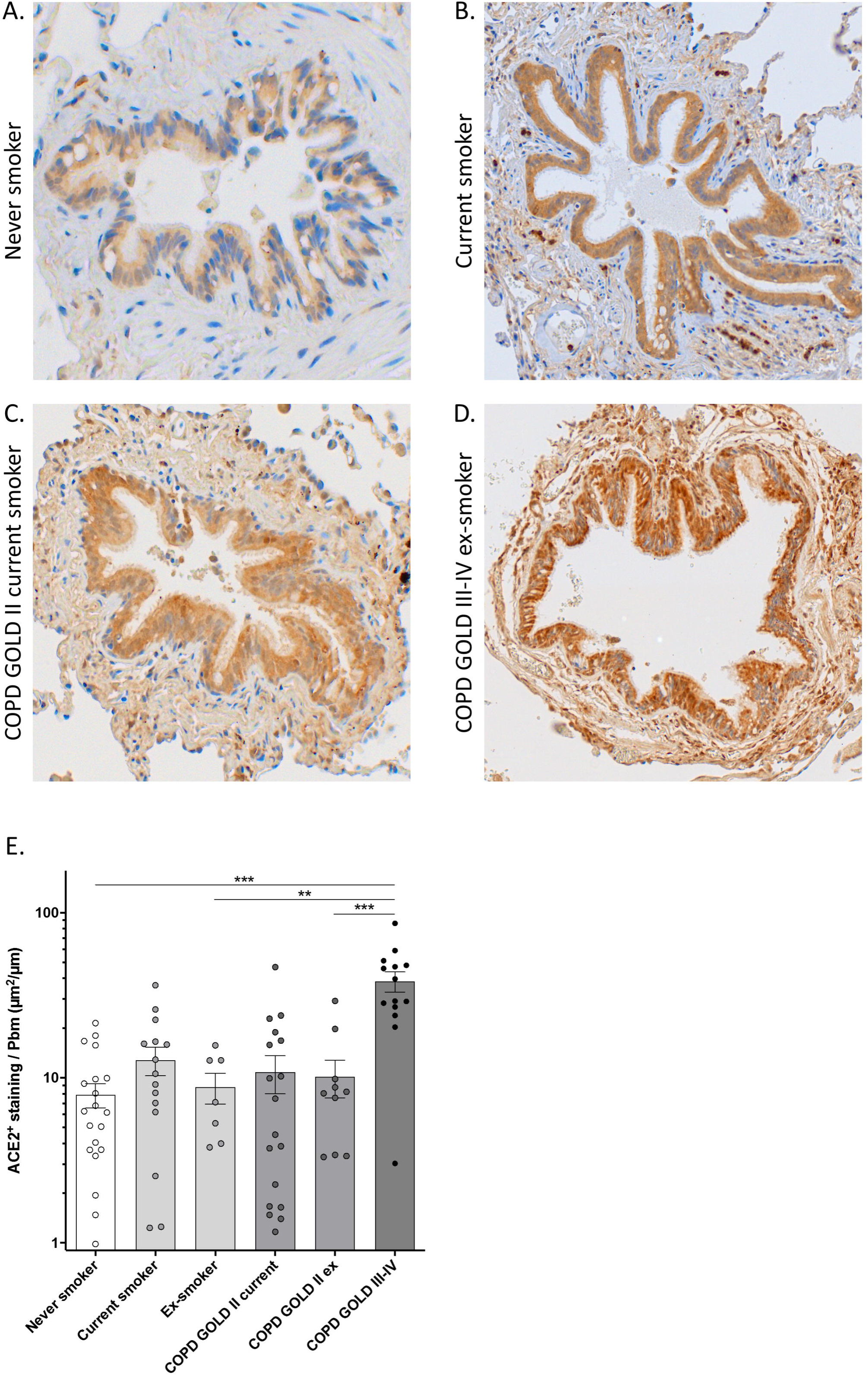
ACE2 protein levels are increased in bronchial epithelium from smokers and COPD subjects. Representative images of ACE2 immunohistochemical staining in bronchial epithelium from **(A)** never smokers, **(B)** current smokers without airflow limitation, **(C)** current smokers with COPD GOLD stage II and **(D)** ex-smokers with COPD GOLD stage III-IV. **(E)** Quantification of ACE2-positive bronchial epithelium using AxioVision software (Zeiss). The area of ACE2-positive signal in each airway was normalized to the length of the basement membrane. Data are presented as means ± SEM. *P<0.05; **P<0.01; ***P<0.001.

### Smoking, COPD, medication use, comorbidities, and lung-specific expression of ACE2

We investigated whether the increased ACE2 levels in subjects with COPD could partially result from cigarette smoking. Although the effect size for the association between COPD and pulmonary ACE2 mRNA expression was affected after adjusting for smoking status in linear regression analyses (unadjusted β 0.28 ± 0.09, p = 0.002; smoke status adjusted β 0.22 ± 0.10, p = 0.036), the association remained statistically significant. Secondly, we tested whether the association between COPD and pulmonary ACE2 mRNA could be confounded by comorbidities or intake of RAAS-inhibitors (ACE-inhibitors or angiotensin receptor blockers (ARB)). Multivariate linear regression analysis demonstrated that current smoking (β 0.20 ± 0.09, p = 0.023) and COPD (β 0.31 ± 0.09, p = 0.001) are both independently associated with increased ACE2 mRNA expression in lung tissue, even after adjustment for covariates including age, sex, diabetes, hypertension and use of RAAS-inhibitors **(Figure 5A)**. Similar findings were observed for the association between COPD and ACE2 protein expression in alveolar tissue **(Figure 5B)** or bronchial epithelium and **(Figure E3)**. Notably, in these models, diabetes mellitus seemed to be significantly associated with increased ACE2 levels in alveolar tissue and bronchial epithelium.

**Figure 5.**
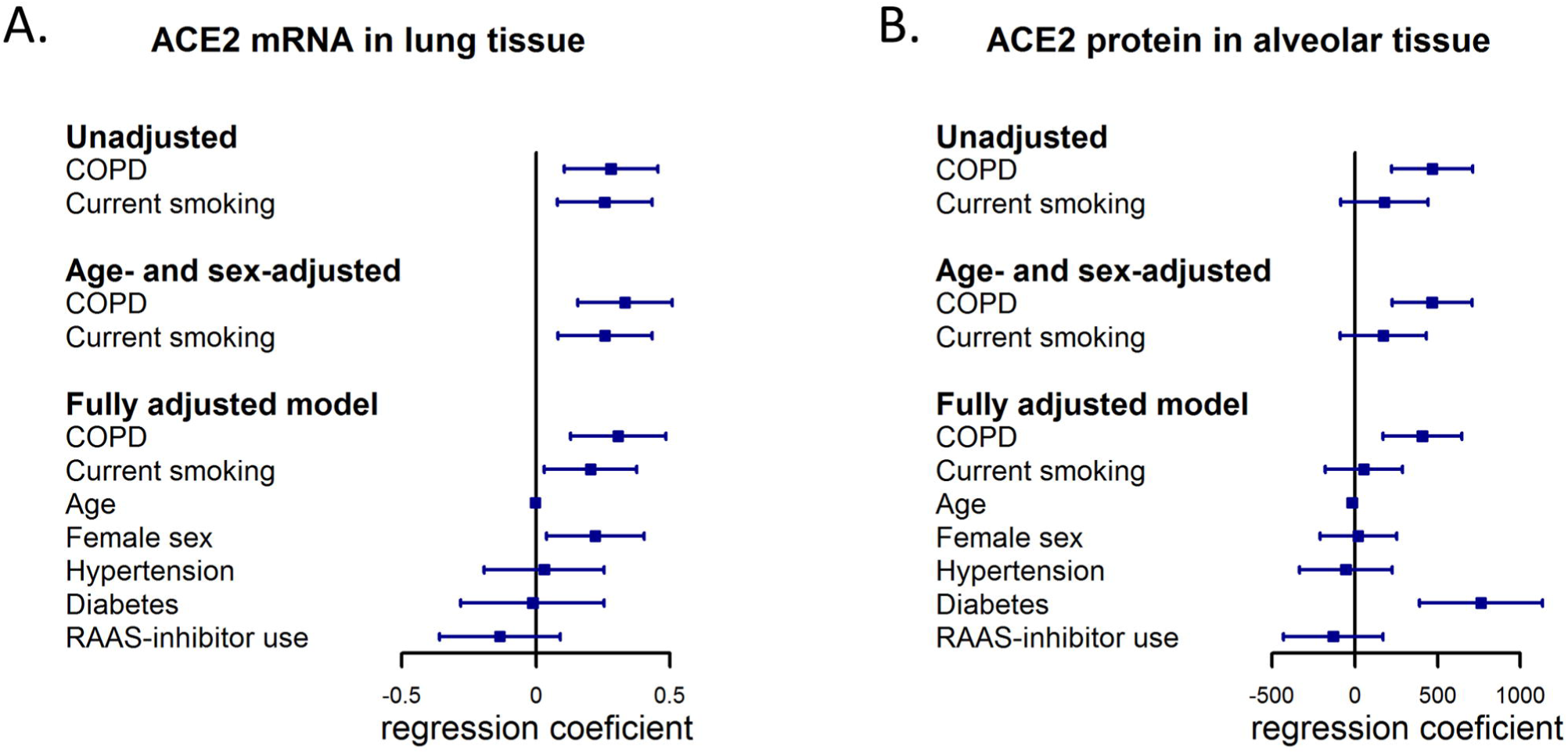
Linear regression analysis of lung-specific expression of ACE2 in smoking, COPD, medication use, and comorbidities. Forrest plots representing linear regression coefficients and 95% confidence intervals of linear regression analysis for **(A)** ACE2 mRNA in lung tissue, and **(B)** ACE2 protein levels in alveolar tissue; in smoking and COPD, and co-variates including age, sex, diabetes, uncontrolled hypertension and ACE-inhibitors or ARBs.

ACE2 mRNA expression was significantly higher in lung tissue of subjects using oral corticosteroid (OCS), but not in lung tissue of subjects using inhaled corticosteroid (ICS) **(Figure E4)**. To test whether increased ACE2 mRNA levels in OCS users may be partially due to underlying COPD, we performed linear regression analyses. Indeed, the significant age- and sex-adjusted association between OCS use and ACE2 mRNA expression (β 0.48 ± 0.16, p < 0.01) did not persist after additional adjustment for COPD (β 0.30 ± 0.17, p = 0.07).

### TMPRSS2 mRNA expression is increased upon cigarette smoke exposure in *in vitro* cultured primary human bronchial epithelial cells (HBECs)

ACE2 and TMPRSS2 mRNA expression was determined by quantitative RT-PCR in HBECs, cultured at air liquid interface (ALI) and exposed to air or cigarette smoke (CS). ACE2 mRNA expression was not altered 3, 6 and 24 hours after CS-exposure **(Figure 6A)**. In contrast, after 6 and 24 hours the TMPRSS2 mRNA expression in CS-exposed HBECs was significantly increased, compared to air exposed cells **(Figure 6B)**.

**Figure 6.**
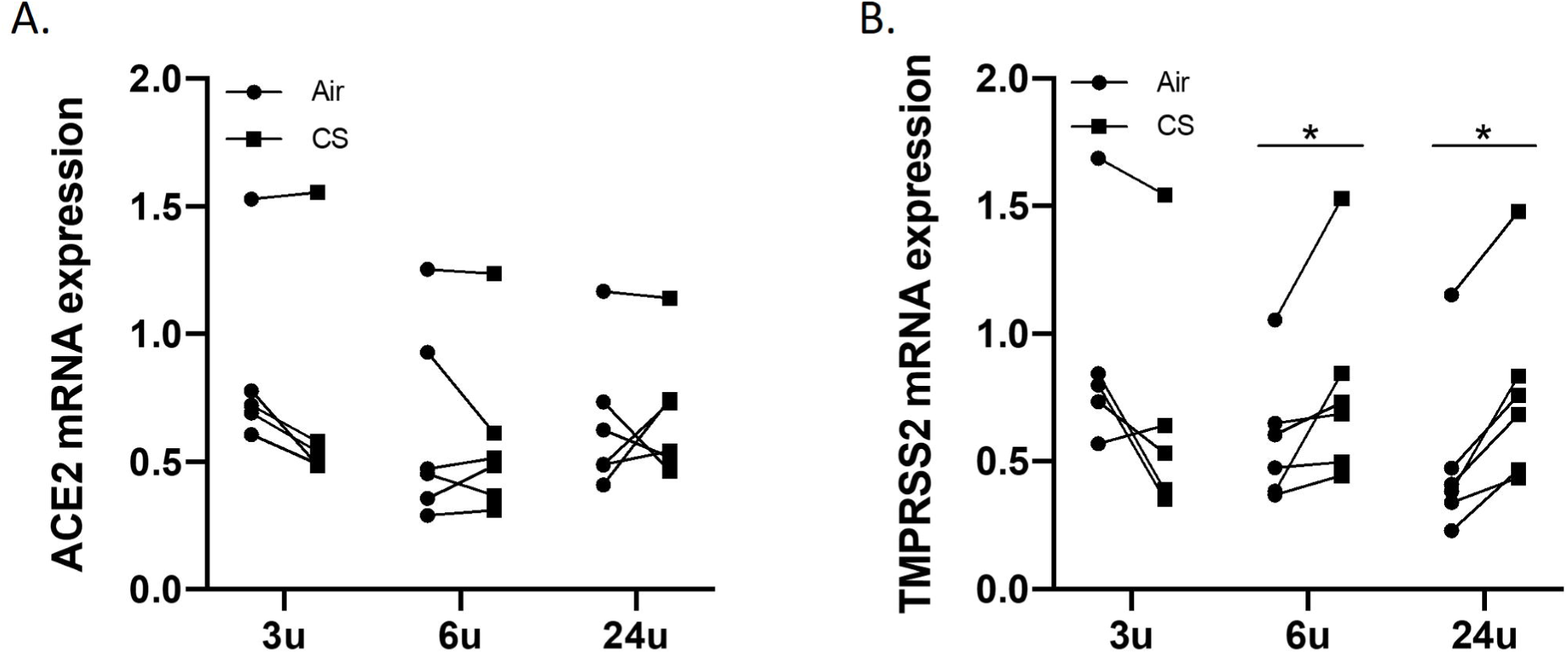
TMPRSS2 mRNA expression is increased upon cigarette smoke exposure in *in vitro* cultured primary human bronchial epithelial cells (HBECs) **(A)** Angiotensinconverting Enzyme-2 (ACE2) and **(B)** Transmembrane protease, serine 2 (TMPRSS2) mRNA expression in HBECs cultured at ALI and exposed to air or cigarette smoke; 3, 6 and 24 hours after the exposures and normalized to the expression of the housekeeping controls glyceraldehyde-3-phosphate dehydrogenase (GAPDH), peptidylprolyl isomerase A (PPIA) and succinate dehydrogenase complex flavoprotein subunit A (SDHA). Data are presented as means ± SEM. *P<0.05; **P<0.01; ***P<0.001.

## DISCUSSION

As healthcare systems around the world are currently under great pressure due to the COVID-19 outbreak, identification of those at high risk is crucial. There is compelling evidence of a more severe course of COVID-19 in smokers and patients with comorbidities such as COPD. We clearly demonstrate an increased pulmonary expression of the SARS-CoV-2 entry receptor ACE2 in smokers and COPD subjects at both mRNA and protein level, by RT-PCR and immunohistochemistry respectively. Our study in smokers with and without COPD confirms our hypothesis that their increased risk for severe COVID-19 may be at least partially attributed to increased ACE2 expression. Importantly, we demonstrate higher ACE2 protein levels not only in bronchial epithelium but also in alveolar epithelium of patients with COPD, which can be directly linked to the site of injury when patients with severe COVID-19 develop dyspnea, hypoxia and pneumonia.

COPD is a highly prevalent disease affecting over 250 million people worldwide. Currently, published data on COVID-19 in patients with COPD are fairly limited [22]. Nevertheless, an increased risk of developing severe COVID-19 as well as a higher mortality, has been reported in patients with COPD and in current smokers [8, 12, 13, 23]. Although there are several possible explanations for the increased susceptibility for severe COVID-19 in patients with COPD, including older age, comorbidities, dysregulated immune defenses and impaired mucociliary clearance, increased pulmonary expression of the SARS-CoV-2 entry receptor ACE2 is most likely an important contributor [24, 25]. Importantly, it has been demonstrated in mouse models that transgenic (over)expression of human ACE2 enhances the pathogenicity of SARS-CoV-1 and SARS-CoV-2 [26, 27]. Moreover, human ACE2 was essential for viral replication in the lung. The same is true for the presence of host cell proteases such as TMPRSS2 and furin, that are essential in the viral entry process [28].

We measured mRNA expression of ACE2 and TMPRSS2 in lung tissue samples from 120 never smokers and smokers with and without airflow limitation and demonstrate higher ACE2 mRNA levels in current smokers and patients with moderate (GOLD II) and severe to very severe COPD (GOLD III-IV). Importantly, ACE2 mRNA expression shows an inverse correlation with physiological parameters of airway obstruction and emphysema. Linear regression analysis confirms that smoking and COPD are associated with increased ACE2 mRNA expression, independent of covariables such as age, gender, comorbidities, and medication use. The association between COPD and ACE2 mRNA seems to be partly driven by smoking as the effect size of the association between COPD and ACE2 mRNA decreased after adjusting for smoking. The remaining statistically significant association between COPD and ACE2 mRNA might be explained by other factors such as aggravated pulmonary and systemic inflammation, previous exacerbations or genetic predisposition. Interestingly, a recent large meta-analysis of transcriptomic data confirms the increased ACE2 mRNA expression in lung tissue of smokers and patients with COPD [28].

TMPRSS2 mRNA expression is only significantly higher in patients with (very) severe COPD. However, there is a significant correlation between TMPRSS mRNA and ACE2 mRNA expression levels, even in sensitivity analyses omitting (very) severe COPD patients from the analysis. Our data suggest that both ACE2 and TMPRSS2 are expressed on the same cells, and co-expression might further increase the chance of viral entry.

By immunohistochemical (IHC) staining we reveal expression of ACE2 in both bronchial and alveolar epithelial cells, with the latter predominantly in alveolar type II (ATII) pneumocytes. Already in 2004 Hamming *et al*. described expression of ACE2 in ATII cells and this has recently been confirmed by both IHC and single cell RNA sequencing analyses [29-31]. Similar analyses also confirm the expression of ACE2 in bronchial epithelial cells, including goblet cells [31, 32].

By quantification of the ACE2 IHC staining, we demonstrate increased protein levels of ACE2 in both alveolar and bronchial epithelium of current smokers and patients with moderate and (very) severe COPD. Linear regression analyses suggest that these associations are partly driven by smoking. Importantly, we are the first to report quantitative evidence for increased ACE2 protein levels in alveolar epithelium, which is the primary site for pathogenic pulmonary infections and could thus explain why smokers and patients with COPD are more prone to develop bilateral pneumonia and/or acute respiratory distress syndrome (ARDS) in severe COVID-19. Indeed, the preferential expression of ACE2 on alveolar type II epithelial cells might make these cells more vulnerable to SARS-CoV-2 infection and virus-induced cell death. Since ATII cells produce surfactants which are crucial for preventing alveolar collapse during breathing, SARS-CoV-2 induced targeting and lysis of ATII cells will alter lung compliance and induce ventilation-perfusion mismatch in severe COVID-19, leading to hypoxia and respiratory failure [33]. Our findings of increased ACE2 levels in bronchial epithelial cells complement the recent paper of Leung *et al*., who reported increased ACE2 mRNA expression in bronchial brushings of smokers and patients with COPD [32].

*In vitro* cultures of primary HBECs at air liquid interface reveal increased mRNA expression of TMPRSS2 6 and 24 hours after a single exposure to cigarette smoke, indicative of an acute effect of smoking on the TMPRSS2 levels in the bronchial epithelium. ACE2 expression might be more dependent on chronic cigarette smoke exposure, since no effect on ACE2 mRNA expression is observed in these short-term *in vitro* experiments.

Inhaled corticosteroids (ICS) are essential in the treatment of asthma and a subgroup of COPD patients (i.e. with a history of frequent exacerbations). Since ICS are often regarded as immunosuppressive, there is a general concern on whether or not to continue the use of ICS during the COVID-19 pandemic. Importantly, our current data shows no association between the ICS use and the expression of ACE2 in the lung. We do report significantly increased ACE2 expression in subjects using oral corticosteroids (OCS), albeit based on a low number of subjects using OCS, implicating the need to confirm these preliminary findings in larger studies. By performing linear regression analyses on our data set, we reveal that lifestyle factors and comorbidities that are linked to a more severe course of COVID-19, such as smoking, COPD, and especially diabetes are associated with a higher expression of ACE2 protein in alveolar tissue. However, due to the observational design of this cross-sectional study, we cannot defer any causal relationships from these associations. Moreover, our results need to be replicated in larger studies and in subjects of other ethnicities.

The main strength of this study is the large number of lung tissue samples from well-phenotyped subjects that are included in the both RT-PCR and IHC analyses. Moreover, IHC allowed us to quantify ACE2 protein levels in both bronchial and alveolar epithelium. However, certain limitations should be kept in mind. First, this study consists mainly of lung tissue from lobectomy. Since these patients might not be representative for the general population, we cannot exclude selection bias. Second, differential information bias may have occurred when questioning smoke status at the time of lobectomy, since COPD status was known to the investigator. However, by carefully checking medical records, we ascertained accurateness of the patient characteristics. Last, although this study consisted of a relatively large and well-phenotyped cohort, samples size may be insufficient for multivariate linear regression analyses. In conclusion, we report higher ACE2 mRNA and protein levels in lung tissue of smokers and subjects with moderate to (very) severe COPD. Importantly, ACE2 protein levels are not only increased in bronchial but also in alveolar epithelium. In addition, TMPRSS2 mRNA expression is higher in lung tissue of (very) severe COPD subjects and increases in *in vitro* cultured HBECs upon acute exposure to cigarette smoke. These results indicate an easier host cell entry of SARS-CoV-2 in both smokers and patients with COPD and are crucial in identifying populations at risk for severe COVID-19.

## Data Availability

All data are available at the department of Respiratory Medicine of the Ghent University Hospital.

## ACKNOWLEDGEMENTS

The authors would like to thank Greet Barbier, Indra De Borle, Katleen De Saedeleer, Anouck Goethals and Ann Neesen, from the Laboratory for Translational Research in Obstructive Pulmonary Diseases, Department of Respiratory Medicine (Ghent University Hospital, Ghent, Belgium) for their excellent technical assistance. Furthermore, we thank Prof. Bart Vanaudenaerde and Dr. Stijn Verleden (Department of Pneumology, Leuven) for providing us with the explant lungs of patients with severe COPD, and Prof. Pieter Hiemstra (Department of Pulmonology, Leiden University Medical Centre) for the cigarette smoke exposure of cultured HBECs.

## Take-Home Message

Increased mRNA and protein levels of ACE2 in alveolar and bronchial epithelium of smokers and patients with COPD might facilitate host cell entry of SARS-CoV-2 and explain their higher risk for a more severe course of COVID-19.

